# Workplace infection control measures and romantic activities of workers during COVID-19 pandemic: a prospective cohort study in Japan

**DOI:** 10.1101/2022.02.13.22270825

**Authors:** Yoshihisa Fujino, Makoto Okawara, Ayako Hino, Keiji Muramatsu, Tomohisa Nagata, Kazunori Ikegami, Seiichiro Tateishi, Mayumi Tsuji, Tomohiro Ishimaru, the CORoNaWork project

## Abstract

**Background:** Due to the COVID-19 pandemic, non-married people are at high risk of loneliness. With social interactions restricted, it is important for non-married people to acquire a new romantic partner for their mental health and quality of life. We hypothesized that infection control efforts in the workplace influence people’s social interactions, including romantic activities.

**Methods:** We conducted an internet-based prospective cohort study from December 2020 (baseline) to December 2021, using self-administered questionnaires. Briefly, 27,036 workers completed the questionnaires at baseline, and when followed up after one year, 18,560 (68.7%) participated. A total of 6,486 non-married individuals with no romantic relationship at baseline were included in the analysis. At baseline they were asked about the implementation of infection control measures in the workplace, and at follow-up they were asked about activities they performed with a view to romantic relationships during the period from baseline to follow-up.

**Results:** Compared to workers in workplaces with no infection control measures, the OR associated with romance-related activities for those in workplaces with seven or more infection control measures was 1.90 (95% CI: 1.45-2.48, p<0.001), and the OR associated with having a new romantic partner was 1.79 (95% CI: 1.20-2.66, p=0.004).

**Conclusions:** Under the COVID-19 pandemic, the implementation of infection control measures in the workplace and the expressed satisfaction with those measures promoted romantic relationships among non-married, single individuals.

## Introduction

Since it started in 2020, the global COVID-19 pandemic has drastically changed people’s daily lives and work styles. As an infection prevention measure, it was recommended that people refrain from or limit normal social activities such as eating out and traveling, and respect physical distance from other people. In Japan, telecommuting was recommended because COVID-19 infections mainly occurred in people of working age, and reducing human contact was deemed important for preventing the spread of infection.^1^ Consequently, whereas 20% of companies in Japan used telecommuting before the COVID-19 pandemic, this figure rose to 57% after the pandemic occurred.^2^

Loneliness is an emerging public health issue resulting from the COVID-19 pandemic situation.^3,4^ Recommendations for physical distance and restrictions on social activity have reduced opportunities for people to interact. In particular, young people, people who are separated or divorced, and those with health issues have reported feeling more lonely during the pandemic.^4^ Loneliness is associated with reduced quality of life, physical health, and mental health.^3–5^

From a public health perspective, it is important that non-married people, a high-risk group for loneliness, are able to establish new romantic relationships, because spending time with family can alleviate loneliness and help maintain good mental well-being.^6^ However, as physical distance is encouraged and social activities continue to be restricted for months or even longer, we considered that it was increasingly difficult for single people to find romantic partners. It is known that the COVID-19 pandemic has impacted romantic relationships in various ways.^7,8^ However, little is known about factors and attributes that influence new romantic activities during the pandemic.

Infection control in the workplace, which is where workers spend most of their time and therefore a major place of human contact, has been reported to affect not only infection prevention but also workers’ mental well-being and performance.^9–12^ COVID-19 measures taken by Japanese employees include refraining from going to work when sick, measuring their body temperature, disinfecting the workplace, and teleworking. However, the extent to which the various control measures are implemented varies with the size of the company, the type of work, and other factors.^13^ Improved infection control measures in the workplace are expected to affect workers’ perception of infection risk and influence their daily behavior and quality of life. Indeed, enhanced infection control measures in the workplace were also reported to be associated with individual preventive behaviors such as hand washing.^10^

We hypothesized that infection control efforts in the workplace also influence people’s interactions, including romantic activities, during the COVID-19 pandemic. It is conceivable that during the pandemic, seeking a new romantic partner is influenced by one’s perception of infection risk. Because people tend to spend more time with new romantic partners and have more physical contact with them, those with greater concerns about infection might be more likely to consider romantic behavior as an infection risk. Furthermore, infection control measures at work might influence the perception of infection risk, including in the context of romantic activities.

Here, we conducted a prospective cohort study to specifically examine whether better infection control in the workplace influences workers’ behaviors related to romantic relationships.

## Methods

The study was conducted under a prospective cohort design between December 2020 and December 2021. Baseline and follow-up surveys were conducted using self-administered questionnaires via the Internet. All participants gave informed consent, and the study was approved by the ethics committee of the University of Occupational and Environmental Health, Japan (reference No. R2-079 and R3-006).

The protocol for the baseline survey has been described elsewhere.^14^ The target population included workers aged 20 to 65 years and in employment at the time of the baseline survey. Sampling was conducted taking into account geographic region, occupation, and sex. Regions were divided into five levels, covering 47 prefectures, according to the level of COVID-19 infection. Occupations were divided into office workers and non-office workers. Thus, a total of 20 blocks of five regions, two occupations, and two sexes was created, and each block was sampled in equal numbers. We planned to collect data from 30,000 people overall, aiming to reach at least 1,500 participants in each block.

The survey was commissioned to Cross Marketing Inc. (Tokyo, Japan). Of their 4.7 million pre-registered monitors, approximately 600,000 were sent an email request to participate in the survey. Of these, 55,045 participated in the initial screening, with 33,302 satisfying the final inclusion criteria. Of those 33,302 participants, 27,036 were included in the analysis, after excluding those judged as submitting untrustworthy responses. The following criteria were used to determine untrustworthy responses: extremely short response time (≤6 minutes), reporting extremely low body weight (<30 kg), reporting extremely short height (<140 cm), inconsistent answers to similar questions throughout the survey (e.g., inconsistency on questions about marital status and area of residence), and wrong answers to a question used solely to identify unreliable responses (“Choose the third largest number from the following five numbers.”).

The follow-up survey was conducted in December 2021, one year after baseline. A total of 18,560 (68.7%) participated in the follow-up. Finally, 6,486 individuals (2,779 men, 3,707 women) who were not married and who were not in a romantic relationship at baseline were included in the analysis. As the study is an internet-based survey, there are no missing values.

### Evaluation of infection control measures in the workplace at baseline

Participants were asked to answer yes or no concerning whether the following measures had been implemented in their workplace: refraining from and restrictions on business trips; refraining from receiving and restrictions on visitors; refraining from meeting or recommending a limit on the number of people at social gatherings and dinners; refraining from or limiting face-to-face internal meetings; wearing masks at all times during working hours; installing partitions and revising the workplace layout; recommending daily temperature checks at home; encouraging telecommuting; prohibiting eating at the work desk; and requesting employees not come to work if feeling unwell.

The questionnaire also asked participants to rate their company’s infection control measures, using the question: “Do you think your company has taken adequate infection control measures for its employees?” Participants responded on a four-point scale: “yes,” “somewhat,” “not really,” “no.”

### Assessment of romantic relationship status

The subjects were asked about their activities in relation to developing romantic relationships during the period from baseline to follow-up (one year). Participants who answered yes to either of the two questions: “Have you taken any action in the past year to find a romantic partner?” “Have you engaged in any marriage activities in the past year?” were classified as “taking action in a romantic relationship”. Those who answered “yes” to the question: “Have you been in a new romantic relationship in the past year?” were classified as “having a new romantic relationship”.

### Other covariates

Information on the subject’s characteristics were collected at the baseline. Participants answered the following questions about themselves in an online form: age, sex, prefecture of residence, marital status (unmarried, bereaved/divorced), job type (mainly desk work, mainly involving interpersonal communication, and mainly labor), number of employees in the workplace, educational background, income, smoking status, and alcohol consumption (6–7 days a week, 4–5 days a week, 2–3 days a week, less than 1 day a week, hardly ever).

### Statistical analyses

Age-sex adjusted odds ratios (ORs) and multivariate adjusted ORs were estimated using a multilevel logistic model nested in the prefecture of residence to take account of regional variability. The ORs of the number of infection control measures in the workplace, and evaluation of those infection control measures associated with “taking action in a romantic relationship” were estimated. The model included age, sex, marital status, job type, income, education, self-rated health, smoking, alcohol drinking, number of employees in the workplace, number of workplace measures against COVID-19, and subjective assessment of whether the company’s infection control measures for employees were adequate. In addition, the incidence rate of COVID-19 by prefecture at baseline was used, and the ORs associated with “having a new romantic relationship” were also estimated.

A *p* value less than 0.05 was considered statistically significant. All analyses were conducted using Stata (Stata Statistical Software: Release 16; StataCorp LLC, TX, USA).

## Results

Table 1 shows the baseline characteristics in relation to the number of infection control measures implemented in the workplace. In workplaces where no infection control measures were taken, 60% of the respondents were men. As the number of infection control measures increased, the percentage of women increased. Compared to workplaces with more infection control measures, workplaces with no infection control measures had lower incomes, more smokers, and were of smaller size.

**Table 1.** Characteristics of subjects according to the number of workplace measures against COVID-19

|  | Number of workplace measures against COVID- 19 |  |  |  |
| --- | --- | --- | --- | --- |
|  | 0<br>n=911 | 1-3<br>n=1383 | 4-6<br>n=1755 | 7-10<br>n=2437 |
| Age, mean (SD) | 47.8 (9.6) | 46.2 (9.7) | 45.0 (9.8) | 45.4 (10.0) |
| Sex, men | 548 (60.2%) | 637 (46.1%) | 691 (39.4%) | 903 (37.1%) |
| Marital status |  |  |  |  |
| Divorce history | 176 (19.3%) | 337 (24.4%) | 381 (21.7%) | 542 (22.2%) |
| Never married | 735 (80.7%) | 1046 (75.6%) | 1374 (78.3%) | 1895 (77.8%) |
| Job type, mainly desk work | 474 (52.0%) | 629 (45.5%) | 788 (44.9%) | 1354 (55.6%) |
| Income (million JPY) |  |  |  |  |
| <300 | 358 (39.3%) | 441 (31.9%) | 424 (24.2%) | 446 (18.3%) |
| 300-499 | 278 (30.5%) | 487 (35.2%) | 641 (36.5%) | 781 (32.0%) |
| 500-799 | 182 (20.0%) | 293 (21.2%) | 455 (25.9%) | 727 (29.8%) |
| >>800 | 93 (10.2%) | 162 (11.7%) | 235 (13.4%) | 483 (19.8%) |
| Education |  |  |  |  |
| Junior high school | 27 (3.0%) | 36 (2.6%) | 18 (1.0%) | 29 (1.2%) |
| High school | 321 (35.2%) | 445 (32.2%) | 465 (26.5%) | 543 (22.3%) |
| Vocational school/college, university, graduate school | 563 (61.8%) | 902 (65.2%) | 1272 (72.5%) | 1865 (76.5%) |
| Self-rated health |  |  |  |  |
| excellent | 38 (2.8%) | 35 (3.7%) | 62 (2.5%) | 43 (2.5%) |
| very good | 103 (7.6%) | 73 (7.7%) | 217 (8.9%) | 151 (8.7%) |
| good | 424 (31.5%) | 304 (32.1%) | 766 (31.3%) | 604 (34.6%) |
| fair | 536 (39.8%) | 376 (39.7%) | 1002 (40.9%) | 706 (40.5%) |
| poor | 247 (18.3%) | 158 (16.7%) | 401 (16.4%) | 240 (13.8%) |
| Current smoker | 276 (30.3%) | 371 (26.8%) | 400 (22.8%) | 503 (20.6%) |
| Alcohol drinking |  |  |  |  |
| 6-7 days per week | 182 (20.0%) | 250 (18.1%) | 243 (13.8%) | 360 (14.8%) |
| 4-5 days per week | 45 (4.9%) | 84 (6.1%) | 100 (5.7%) | 155 (6.4%) |
| 2-3 days per week | 94 (10.3%) | 139 (10.1%) | 215 (12.3%) | 247 (10.1%) |
| Less than 1 day per week | 130 (14.3%) | 234 (16.9%) | 328 (18.7%) | 461 (18.9%) |
| Hardly ever | 460 (50.5%) | 676 (48.9%) | 869 (49.5%) | 1214 (49.8%) |
| Number of employees in the workplace |  |  |  |  |
| 1-4 | 511 (56.1%) | 398 (28.8%) | 227 (12.9%) | 178 (7.3%) |
| 5-49 | 212 (23.3%) | 499 (36.1%) | 485 (27.6%) | 315 (12.9%) |
| 50-499 | 109 (12.0%) | 331 (23.9%) | 556 (31.7%) | 817 (33.5%) |
| >>500 | 79 (8.7%) | 155 (11.2%) | 487 (27.7%) | 1127 (46.2%) |
| “Do you think your company has taken adequate infection control measures for its employees?” |  |  |  |  |
| Yes | 79 (8.7%) | 113 (8.2%) | 227 (12.9%) | 560 (23.0%) |
| Somewhat | 314 (34.5%) | 659 (47.7%) | 1036 (59.0%) | 1501 (61.6%) |
| Not really | 221 (24.3%) | 367 (26.5%) | 362 (20.6%) | 292 (12.0%) |
| No | 297 (32.6%) | 244 (17.6%) | 130 (7.4%) | 84 (3.4%) |
| New romantic relationships | 70 (7.7%) | 160 (11.6%) | 239 (13.6%) | 364 (14.9%) |

Table 2 shows the ORs for the number of infection control measures and activities related to romantic relationships. Compared to workers in workplaces with no infection control measures, those in workplaces with seven or more measures were more likely to engage in romantic relationship-related activities (OR=1.84, 95% CI: 1.42-2.47, p<0.001). The multivariate adjusted model also showed this association (OR=1.90, 95% CI: 1.45-2.48, p<0.001). We also found a linear relationship between the number of infection control measures and activities in a romantic relationship (p for trend < 0.001).

**Table 2.** Association between number of workplace measures against COVID-19 and activities in a new romantic relationship

|  | age-adjusted |  |  |  | multivariate* |  |  |  |
| --- | --- | --- | --- | --- | --- | --- | --- | --- |
|  | OR | 95% CI |  | p | OR | 95% CI |  | p |
| Number of workplace measures against COVID-19 |  |  |  |  |  |  |  |  |
| 0 | reference |  |  |  | reference |  |  |  |
| 1-3 | 1.46 | 1.08 | 1.97 | 0.015 | 1.43 | 1.07 | 1.90 | 0.016 |
| 4-6 | 1.63 | 1.22 | 2.17 | 0.001 | 1.53 | 1.19 | 1.97 | 0.001 |
| 7-10 | 1.87 | 1.42 | 2.47 | <0.001 | 1.90 | 1.45 | 2.48 | <0.001 |
|  |  |  |  | <0.001<sup>†</sup> |  |  |  | <0.001<sup>†</sup> |
| My company has adequate infection control measures in place for its employees. |  |  |  |  |  |  |  |  |
| Strongly disagree | reference |  |  |  | reference |  |  |  |
| Disagree | 1.15 | 0.85 | 1.56 | 0.362 | 1.12 | 0.82 | 1.53 | 0.481 |
| Agree | 1.28 | 0.98 | 1.68 | 0.069 | 1.25 | 0.95 | 1.65 | 0.116 |
| Strongly agree | 1.68 | 1.24 | 2.28 | 0.001 | 1.55 | 1.13 | 2.12 | 0.007 |
|  |  |  |  | <0.001<sup>†</sup> |  |  |  | 0.003<sup>†</sup> |

\* The model included age, sex, marital status, job type, income, education, self-rated health, smoking, alcohol drinking, number of employees in the workplace, number of workplace measures against COVID-19, and subjective assessment of whether the company has adequate infection control measures in place for employees.
<sup>†</sup> p for trend

Similarly, workers who perceived their company’s infection control self-perception as adequate compared to those who considered it inadequate were more likely to engage in romantic relationship-related activities (OR=1.68, 95% CI:1.24-2.28, p=0.001). This relationship was similar in the multivariate model (OR=1.55, 95% CI:1.13-2.12, p=0.007). Furthermore, workers’ perception of infection control in the workplace showed a linear relationship in the order of strongly disagree, disagree, agree, and strongly disagree (p for trend =0.003).

Table 3 shows the ORs of participants who actually established a romantic relationship during the period between baseline and follow-up. The more infection control measures were implemented in the workplace, the more people started to have new romantic relationships. In the multivariate analysis, the OR for those who reported seven or more measures in the workplace compared to those who reported that no infection control measures existed was 1.79 (95% CI: 1.20-2.66, p=0.004). There was also a linear relationship: the higher the number of measures, the greater the likelihood of starting a new romantic relationship (p for trend <0.001). In addition, there was a nonsignificant tendency (p for trend <0.081) for those who felt that the level of infection control in the workplace was adequate to establish a new romantic relationship.

**Table 3.** Association between number of workplace measures against COVID-19 and having a new romantic relationship

|  | age-adjusted |  |  |  | multivariate* |  |  |  |
| --- | --- | --- | --- | --- | --- | --- | --- | --- |
|  | OR | 95% CI |  | p | OR | 95% CI |  | p |
| Number of workplace measures against COVID-19 |  |  |  |  |  |  |  |  |
| 0 | reference |  |  |  | reference |  |  |  |
| 1-3 | 1.22 | 0.82 | 1.83 | 0.327 | 1.11 | 0.73 | 1.67 | 0.631 |
| 4-6 | 1.25 | 0.85 | 1.83 | 0.265 | 1.15 | 0.76 | 1.72 | 0.511 |
| 7-10 | 1.84 | 1.28 | 2.64 | 0.001 | 1.79 | 1.20 | 2.66 | 0.004 |
|  |  |  |  | <0.001<sup>†</sup> |  |  |  | <0.001<sup>†</sup> |
| My company has adequate infection control measures in place for its employees. |  |  |  |  |  |  |  |  |
| Strongly disagree | reference |  |  |  | reference |  |  |  |
| Disagree | 1.36 | 0.89 | 2.06 | 0.156 | 1.30 | 0.85 | 2.00 | 0.224 |
| Agree | 1.40 | 0.96 | 2.05 | 0.079 | 1.32 | 0.90 | 1.95 | 0.156 |
| Strongly agree | 1.78 | 1.16 | 2.71 | 0.008 | 1.53 | 0.99 | 2.36 | 0.057 |
|  |  |  |  | 0.011<sup>†</sup> |  |  |  | 0.081<sup>†</sup> |

\* The model included age, sex, marital status, job type, income, education, self-rated health, smoking, alcohol drinking, number of employees in the workplace, number of workplace measures against COVID-19, and subjective assessment of whether the company has adequate infection control measures in place for employees.
<sup>†</sup> p for trend

## Discussion

This study showed that greater implementation of infection control measures in the workplace is associated with more activities toward initiating romantic relationships. Furthermore, workers who expressed satisfaction with their company’s infection control measures were more likely to be active in initiating a relationship than those who felt that their company’s measures were inadequate. These results support the hypothesis that infection control efforts in the workplace influence workers’ romantic activities during the COVID-19 pandemic.

Pietromonaco et al. proposed a conceptual framework for the impact of the COVID-19 pandemic on romantic relationships, based on the adaptive process from the perspective of relationship science.^8^ The adaptive process involve interaction in which couples respond to external stressors and difficulties by mutually supporting each other and functioning through their problems.^8,15^ When subjected to an external stressor, COVID-19 being a good example, couples that have existing problems will experience a decline in their relationship. The co-presence of emotional factors such as anxiety and depression also results in a lack of supportive affection when couples need to support each other, resulting in negative interactions.^16,17^ A positive association between adequate infection control measures in the workplace and workers’ mental health and anxiety has been reported.^12^ Infection control in the workplace may have a moderating effect on workers’ feelings of vulnerability by alleviating anxiety, and thereby positively influence within-couple relationships.

Adequate infection control measures in the workplace may act as a deterrent to self-regulatory depletion in workers and promote the initiation and construction of romantic relationships. The theory of self-regulatory depletion has been proposed as a mechanism by which external stress can lead to decreased cooperation and satisfaction among partners.^18^ Because coping with external stress requires individual effort, it depletes self-regulatory capacity, leading to more negative behavior toward partners and inhibiting dyadic relationships.^15,18^ Couples experiencing increased daily stress may show more criticism of their partner, which has been attributed to self-regulation depletion.^18^ In the COVID-19 pandemic, various external stresses including economic problems, loneliness, employment instability, physical limitations, and limited social activities can make it difficult for couples to function in a complementary and supportive manner, and diminish romantic relationships. Working under a high perceived risk of infection can lead to negative attitudes toward one’s partner or potential partner and a decline in the relationship, as one’s self-regulatory capacity is depleted because of the effort required to avoid crisis. Adequate infection control measures in the workplace can alleviate the external stress and anxiety that COVID-19 brings to couples, including the anxiety that infection may be introduced to couples and families; they can help maintain good dyadic adjustment and psychological well-being, and thereby the quality of the relationship.

We chose socioeconomic factors related to marriage as confounding factors in this study because romantic activities are in some ways similar to those of marriage, albeit that the sociodemographic drivers of romantic activities are not clear. Even after adjusting for factors such as age, income, and health status, we found an association between workplace infection control and romantic activity. This result implies that the mechanisms underlying the association of adequate workplace infection control with romantic activity may depend on factors other than socioeconomic ones. It has been reported that infection control in the workplace is associated with workers’ risk perception, individual preventive behavior, mental health, and stress.^10,12,19^ It seems likely that these factors can either promote or retard the intention to engage in romantic behavior. Nevertheless, this question warrants further examination.

Some limitations of this study can be mentioned. First, although we assessed romantic activities using self-reports, there is little reason to doubt participants’ recollections about whether or not they started a romantic relationship. Also, as the study involved an anonymous survey via the Internet, there was little motivation for false reporting. Second, our operational definition of romantic activity might be questioned. The presence or absence of a sexual relationship, homosexuality, or multiple partners are unknown. However, we believe that any impact of not distinguishing between these factors was minimal in the context of this study. Third, it is not clear how participants found their romantic partners, whether at parties, or by social networking applications, for example. It is possible that social networking activities have become more prevalent during the COVID-19 pandemic,^20–22^ but any impact on this study remains unknown.

In conclusion, under the COVID-19 pandemic, implementation of infection control measures in the workplace and the degree of satisfaction with these measures promoted romantic relationships among non-married individuals who did not have a romantic partner. We propose that infection control measures at work facilitate romantic activities and the establishment of romantic relationships by alleviating anxiety and supplementing self-regulatory capacity depleted due to the external stress of COVID-19. Single people are at high risk for loneliness, and the latter has intensified during the COVID-19 pandemic. Meeting and establishing a relationship with a romantic partner may be an important factor in maintaining mental well-being, for as long as physical distancing continues to be recommended.

## Data Availability

Data cannot be shared for ethical reasons.

## Acknowledgements

The authors declare they have no conflict of interest with respect to this study and article.

The study was supported and partly funded by a research grant from the University of Occupational and Environmental Health, Japan (no grant number); Japanese Ministry of Health, Labour and Welfare (H30-josei-ippan-002, H30-roudou-ippan-007, 19JA1004, 20JA1006, 210301-1, and 20HB1004); Anshin Zaidan (no grant number), the Collabo-Health Study Group (no grant number), and Hitachi Systems, Ltd. (no grant number) and scholarship donations from Chugai Pharmaceutical Co., Ltd. (no grant number). The funder was not involved in the study design, collection, analysis, interpretation of data, the writing of this article or the decision to submit it for publication.

The current members of the CORoNaWork Project, in alphabetical order, are as follows Dr. Hajime Ando, Dr. Hisashi Eguchi, Dr. Yoshihisa Fujino (present chairperson of the study group), Dr. Arisa Harada, Dr. Ayako Hino, Dr. Kazunori Ikegami, Dr. Tomohiro Ishimaru, Dr. Kyoko Kitagawa, Ms. Ning Liu, Dr. Kosuke Mafune, Dr. Shinya Matsuda, Dr. Ryutaro Matsugaki, Dr. Koji Mori, Dr. Keiji Muramatsu, Dr. Masako Nagata, Dr. Tomohisa Nagata, Dr. Akira Ogami, Dr. Makoto Okawara, Dr. Rie Tanaka, Dr. Seiichiro Tateishi, Dr. Kei Tokutsu, and Dr. Mayumi Tsuji. All members are affiliated with the University of Occupational and Environmental Health, Japan.

## Data Availability

Data cannot be shared for ethical reasons.

## References

1. Ministry of health labour and welfare, Japan. Visualizing the data: information on COVID-19 infections. Accessed February 1, 2022. https://covid19.mhlw.go.jp/en/

2. Tokyo Metropolitan Government. [Results of a survey on telework adoption rates]. Accessed February 1, 2022. https://www.metro.tokyo.lg.jp/tosei/hodohappyo/press/2021/01/22/17.html

3. Pai N, Vella SL. COVID-19 and loneliness: A rapid systematic review. Aust N Z J Psychiatry. 2021;55(12):1144–1156.

4. Groarke JM, Berry E, Graham-Wisener L, McKenna-Plumley PE, McGlinchey E, Armour C. Loneliness in the UK during the COVID-19 pandemic: Cross-sectional results from the COVID-19 Psychological Wellbeing Study. PLoS ONE. 2020;15(9):e0239698.

5. Konno Y, Nagata M, Hino A, et al. Association between loneliness and psychological distress: A cross-sectional study among Japanese workers during the COVID-19 pandemic. Prev Med Rep. 2021;24:101621.

6. Fujii R, Konno Y, Tateishi S, et al. Association between time spent with family and loneliness among Japanese workers during the COVID-19 pandemic: A cross-sectional study. Front Psychiatry. 2021;12:786400.

7. Weber DM, Wojda AK, Carrino EA, Baucom DH. Love in the time of COVID-19: A brief report on relationship and individual functioning among committed couples in the United States while under shelter-in-place orders. Fam Process. Published online July 27, 2021. doi:10.1111/famp.12700

8. Pietromonaco PR, Overall NC. Applying relationship science to evaluate how the COVID-19 pandemic may impact couples’ relationships. Am Psychol. 2020;76(3):438–450.

9. Sasaki N, Kuroda R, Tsuno K, Kawakami N. Workplace responses to COVID-19 associated with mental health and work performance of employees in Japan. J Occup Health. 2020;62(1):e12134.

10. Kawasumi M, Nagata T, Ando H, et al. Association between preventive measures against workplace infection and preventive behavior against personal infection. Ind Health. Published online November 16, 2021. doi:10.2486/indhealth.2021-0162

11. Kurogi K, Ikegami K, Eguchi H, et al. A cross-sectional study on perceived workplace health support and health-related quality of life. J Occup Health. 2021;63(1):e12302.

12. Yasuda Y, Ishimaru T, Nagata M, et al. A cross-sectional study of infection control measures against COVID-19 and psychological distress among Japanese workers. J Occup Health. 2021;63(1):e12259.

13. Ishimaru T, Nagata M, Hino A, et al. Workplace measures against COVID-19 during the winter third wave in Japan: Company size-based differences. J Occup Health. 2021;63(1):e12224.

14. Fujino Y, Ishimaru T, Eguchi H, et al. Protocol for a nationwide internet-based health survey of workers during the COVID-19 pandemic in 2020. J UOEH. 2021;43(2):217–225.

15. Karney BR, Bradbury TN. The longitudinal course of marital quality and stability: A review of theory, methods, and research. Psychol Bull. 1995;118(1):3–34.

16. Davila J, Bradbury TN, Cohan CL, Tochluk S. Marital functioning and depressive symptoms: Evidence for a stress generation model. J Pers Soc Psychol. 1997;73(4):849–861.

17. Beck LA, Pietromonaco PR, DeBuse CJ, Powers SI, Sayer AG. Spouses’ attachment pairings predict neuroendocrine, behavioral, and psychological responses to marital conflict. J Pers Soc Psychol. 2013;105(3):388–424.

18. Buck AA, Neff LA. Stress spillover in early marriage: the role of self-regulatory depletion. J Fam Psychol. 2012;26(5):698–708.

19. Smith PM, Oudyk J, Potter G, Mustard C. Labour market attachment, workplace infection control procedures and mental health: A cross-sectional survey of Canadian non-healthcare workers during the COVID-19 pandemic. Ann Work Expo Health. 2021;65(3):266–276.

20. Gibson AF. Exploring the impact of COVID-19 on mobile dating: Critical avenues for research. Soc Personal Psychol Compass. 2021;15(11):e12643.

21. Portolan L, McAlister J. Jagged love: Narratives of romance on dating apps during COVID-19. Sex Cult. Published online July 20, 2021:1–19.

22. Wiederhold BK. How COVID has changed online dating-and what lies ahead. Cyberpsychol Behav Soc Netw. 2021;24(7):435–436.

